# Promise vs. Proof in Digital Interventions for Antimicrobial Stewardship: A Systematic Review and Meta-Analysis of Randomized Controlled Trials

**DOI:** 10.64898/2026.06.01.26354656

**Authors:** Ana Paula Matos Porto, Marcela Símaro Gomes, Vítor Falcão de Oliveira, Herman Mwanja, Nina Zhu, Alison Holmes, Anna S. Levin, Silvia Figueiredo Costa, the CAMO-Net Brazil Study Group

**Author notes:** Corresponding authors: /fax (email).

## Abstract

**Background:** Digital antimicrobial stewardship (AMS) interventions, such as clinical decision support systems, audit-and-feedback platforms, and electronic prescribing tools, have been increasingly adopted to improve antibiotic use. However, the effectiveness of these interventions across healthcare settings remains uncertain, and the certainty of the evidence has not been comprehensively evaluated. The objective of this study was to provide a comprehensive understanding of the role of digital interventions in optimizing antimicrobial use and improving clinical outcomes within a broad spectrum of healthcare settings.

**Methods:** We conducted a systematic review and meta-analysis of randomized controlled trials evaluating digital AMS interventions that followed PRISMA 2020 guidelines and registered in PROSPERO (CRD420251178854) and funded by the Wellcome Trust CAMO-Net programme. Searches were performed across major databases. Primary outcomes included the appropriateness of antibiotic prescriptions and the antibiotic prescription rate. Secondary outcomes included 30-day mortality, 30-day hospital readmission, and length of hospital stay (LOS). Random-effects models were used to pool effect sizes. Risk of bias was assessed using RoB 2.0, and certainty of evidence was rated using GRADE. A Summary of Findings table was prepared to present effect estimates, sample sizes, and evidence certainty.

**Results:** Eleven RCTs met the inclusion criteria, and nine were included in the quantitative synthesis. Digital AMS interventions did not show a significant effect on appropriateness of antibiotic prescribing (RR 0.99, 95%CI 0.93–1.05; very low certainty). There was no reduction in antibiotic prescription (RR 0.98, 95%CI 0.88–1.09), with substantial statistical heterogeneity (I² = 71%) and very low certainty. Across clinical outcomes, digital AMS showed no effect on 30-day mortality (RR 0.91, 95%CI 0.77–1.09; very low certainty) or 30-day readmission (RR 0.95, 95%CI 0.79–1.14; very low certainty). For LOS, results were inconsistent across studies, and the pooled effect showed no clinically meaningful change (MD 0.17 days, 95%CI –0.01 to 0.35; very low certainty). Most trials had “some concerns” of bias due to deviations from intended interventions.

**Conclusion:** Meta-analyses of digital AMS RCTs showed a lack of evidence with a high level of certainty on antibiotic prescribing or clinical outcomes due to high heterogeneity in interventions and study designs, as well as RCTs’ limitations (no adoption/fidelity metrics).

## Introduction

Effective antimicrobial stewardship (AMS) programs optimize antimicrobial prescribing, a cornerstone against AMR via judicious use, yet direct resistance reductions lack robust evidence.^1^ Therefore, proceed cautiously on resistance claims.^2,3^ These programs typically involve coordinated actions by a multidisciplinary team, co-led by an infectious disease (ID) physician and a clinical pharmacist, who oversee adherence to guidelines, de-escalation protocols, and prospective audits. However, in lower- and middle-income countries (LMICs) and rural areas of high-income countries (HICs), critical components for successful AMS implementation are often absent or underdeveloped due to shortages in human resources (e.g., limited ID specialists), inadequate infrastructure, and financial constraints.^4^

The ID specialist shortage underscores the urgent need for innovative solutions, such as digital health interventions, defined as a discrete function of digital health technology providing a health objective.^5^ For AMS purposes, this may include computer-based applications, mobile apps, eHealth platforms, and clinical decision support systems (CDSS), which can democratize access to expert guidance in remote settings. These tools usually allow leveraging real-time data integration, algorithmic guidance, and remote consultation features to address common AMS challenges, such as overprescribing due to diagnostic uncertainty.^6^

Despite the growing proliferation of digital tools within AMS with a significant acceleration over the past five years, building upon a decade of foundational development, particularly since the onset of the COVID-19 pandemic, robust evidence regarding their effectiveness in real-world clinical settings remains notably limited, mostly from observational studies.^7^ Building upon this identified gap, this review endeavors to critically analyze and synthesize findings exclusively from RCTs, supposed to be more methodologically robust, seeking to provide a comprehensive understanding of the role of digital interventions in optimizing antimicrobial use and improving clinical outcomes within a broad spectrum of healthcare settings.

## Methods

We conducted a systematic literature review in accordance with the recommendations of the Preferred Reporting Items for Systematic Reviews and Meta-Analyses (PRISMA).^8^ We registered the protocol on PROSPERO (CRD420251178854) prior to data extraction.

### Search strategy

We conducted the search on August 22nd, 2025, in the following databases: PubMed/Medline, BVS/LILACS, Scopus, and Embase. We used no filters for language or date of publication.

A comprehensive search strategy was developed in consultation with a specialist librarian at the University of São Paulo. The review involved evaluating RTCs that assessed digital interventions or included these digital tools as part of antimicrobial stewardship interventions in all healthcare settings (primary care, acute hospitals, and long-term care facilities). The query expression was a combination of at least 2 terms: one related to antimicrobial stewardship (AMS) (“antimicrobial stewardship” or “antibiotic stewardship” or “stewardship, antibiotic” or “stewardship, antimicrobial”), and another to the intervention (“digital tool” or “computer” or “mobile” or “electronic” or “eHealth” or “Telemedicine”). The complete list of search terms is provided in Supplementary Appendix 1.

### Eligibility and study selection

Eligible articles explicitly described the use of digital interventions to enhance antimicrobial stewardship, guiding prescribing or providing actionable stewardship support, including computer-based audit-and-feedback systems, mobile applications (mHealth), eHealth platforms, and clinical decision support systems (CDSS) in healthcare settings. We excluded studies that focused solely on the use of automatic general alerts integrated into the EMR without personalized recommendation input, justified by the well-known phenomenon of alert fatigue.^9^ We also excluded studies based on the use of patient-facing applications unrelated to AMS decision-making. Furthermore, we excluded studies with a non-randomized controlled trial (Non-RCT) design as described in Figure 1. We chose to include only RCTs, as they represent the highest level of evidence for evaluating interventions.

**Figure 1.**
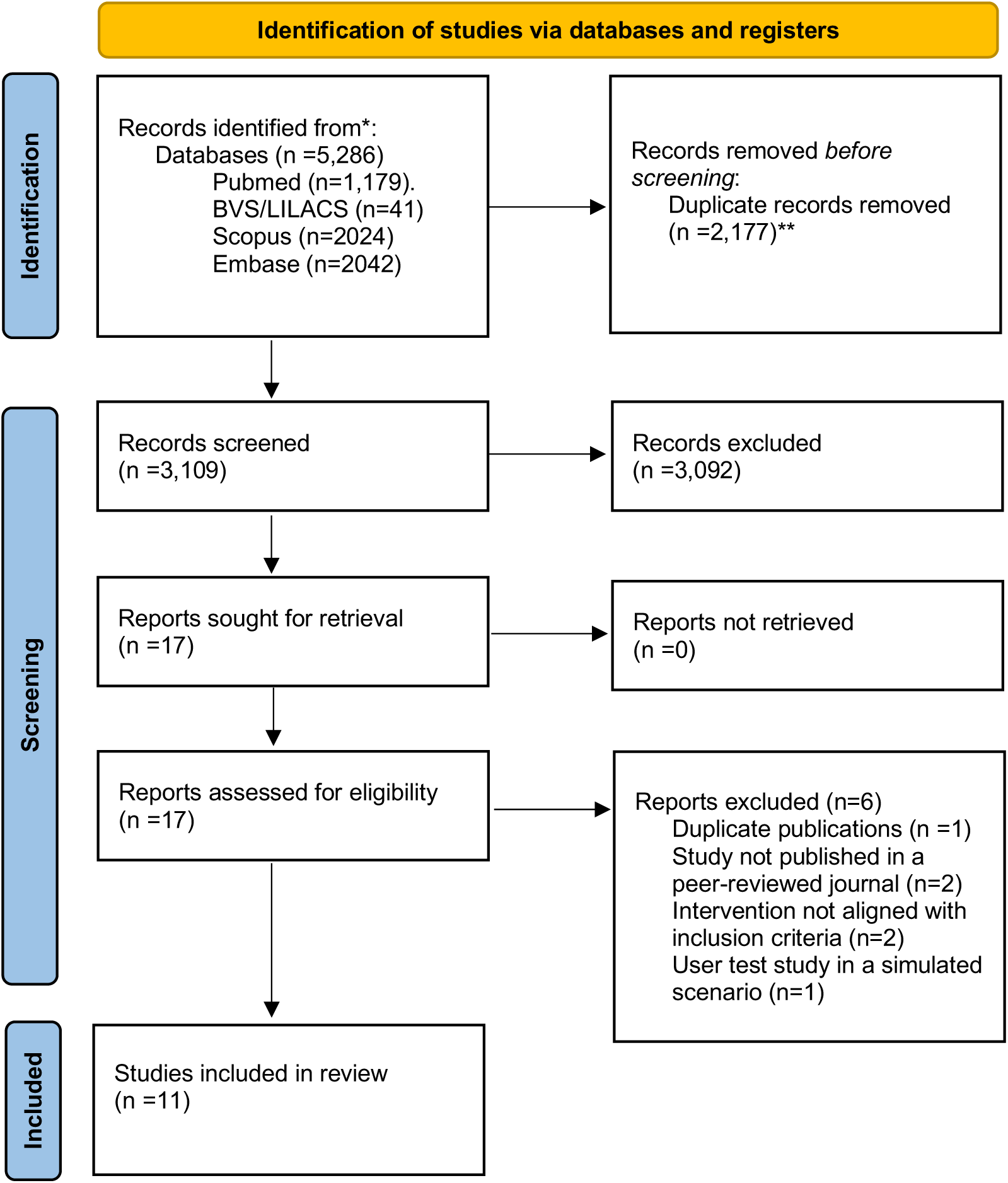
PRISMA flow diagram for meta-analysis of digital interventions for antimicrobial stewardship. *Number of articles disaggregated per database **Covidence used to remove duplicates

We first performed an initial screening of titles and abstracts to assess potential relevance (done independently by APMP and MSG). Afterwards, we obtained full-text articles, re-evaluated their eligibility, and determined their final inclusion or exclusion (APMP and MSG, independently). Disagreement was solved by consensus.

### Data extraction

Data extraction was carried out by two independent investigators (APMP and MSG). Disagreements were resolved through consensus.

The following data were extracted: author name, country of the study, year of publication, study setting, sample size, type of randomization, unit of analysis (individual or cluster), patient population (adult or pediatric), type of digital intervention, multimodal strategy, infection site, control group, outcomes, and analysis methods (intention-to-treat or per-protocol).

Primary outcomes were appropriateness of antibiotic prescribing and antibiotic prescription rate, with their specific definitions detailed in the supplementary material as per the selected studies.

The secondary outcomes were: 30-day mortality, length of stay in days, and 30-day hospital readmission. While cost and antimicrobial resistance were predefined in the protocol, they were subsequently excluded from the analysis as a minimum of two studies per outcome was required for quantitative synthesis.

### Risk of bias

The articles were evaluated for risk of bias using the Cochrane Risk of Bias tool.^10^ Two authors (MSG and APMP) reviewed the risk of bias of each study.

### Certainty of evidence

The GRADE (Grading of Recommendations, Assessment, Development and Evaluation) approach was used to assess the certainty of outcome evidence.^11^

### Data analysis

We performed the analysis using the RStudio software version 1.4 using package meta. Meta-analyses were used to generate summary relative risk estimates and 95% confidence intervals (CI). Random-effects models were used due to high heterogeneity. Summary associations were interpreted as statistically significant (p < 0.05) if the 95% CIs did not include the value of 1.0 in their range. Statistical heterogeneity was measured using I².

## Results

### Study selection

We identified 5,286 articles through database searches, of which 2,177 duplicates were removed. Following title and abstract screening of the 3,109 articles, 3,092 articles were removed. The remaining 17 full-text articles were assessed for eligibility, and six were excluded with reasons (Figure 1). Ultimately, 11 articles that documented RCTs^12–22^ published between 2019 and March 2025, were included in the review.

### Study characteristics

The 11 included randomized controlled trials collectively enrolled a total of 764,377 participants. Most articles reported RCTs that were conducted in high-income countries, predominantly in the USA (6/11) and Europe (4/11), with most published from 2022 onward (Table S1). Study settings included inpatient hospitals (n=5), emergency departments (n=2), a nursing home (n=1), and primary care, including one telemedicine-based service (n=3).

Among the 11 included trials, five enrolled adult populations, four pediatric populations, and two mixed or general populations. Respiratory tract infections were the most commonly studied condition (5), followed by urinary tract infections (3). Clinical decision support systems (CDSS) were the predominant digital intervention, used in 8 of the studies (Table S1).

Two studies were excluded from the quantitative synthesis due to outcome incompatibility or insufficient data. Of the nine studies synthesized, one was judged to be at high risk of bias and five to have some concerns, primarily due to deviations from intended interventions (RoB 2.0 Domain 2) related to period and carryover effects in crossover trials (Figure 2).

**Figure 2:**
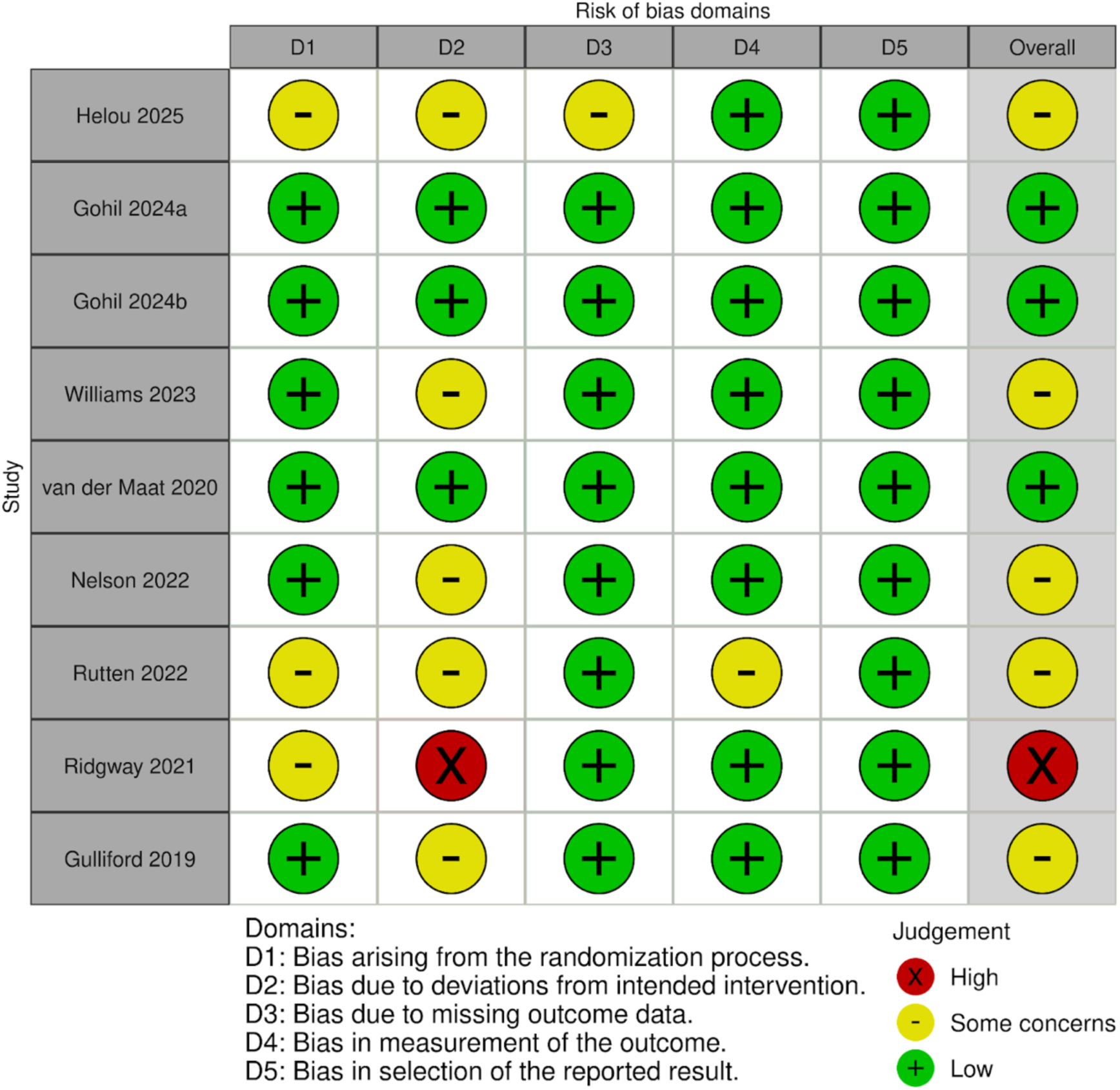
Meta-analysis of randomised controlled studies evaluating the use of digital interventions in antimicrobial stewardship: results of the risk of bias assessment for each study. Scores are shown for each criterion.

Publication bias could not be formally assessed using funnel plots because each outcome included a limited number of studies (2–5), which was insufficient for reliable interpretation.

### Primary outcomes

The pooled rate of appropriateness of antibiotic prescribing (RR: 0.99, 95%CI: 0.93 - 1.05; p-value =0.14; I² = 49%) and antibiotic prescription rate did not demonstrate a significant difference between digital intervention and control groups (RR: 0.98, 95%CI: 0.88 - 1.09; p-value < 0.01; I² = 71) (Figure 3A and 4B).

**Figure 3:**
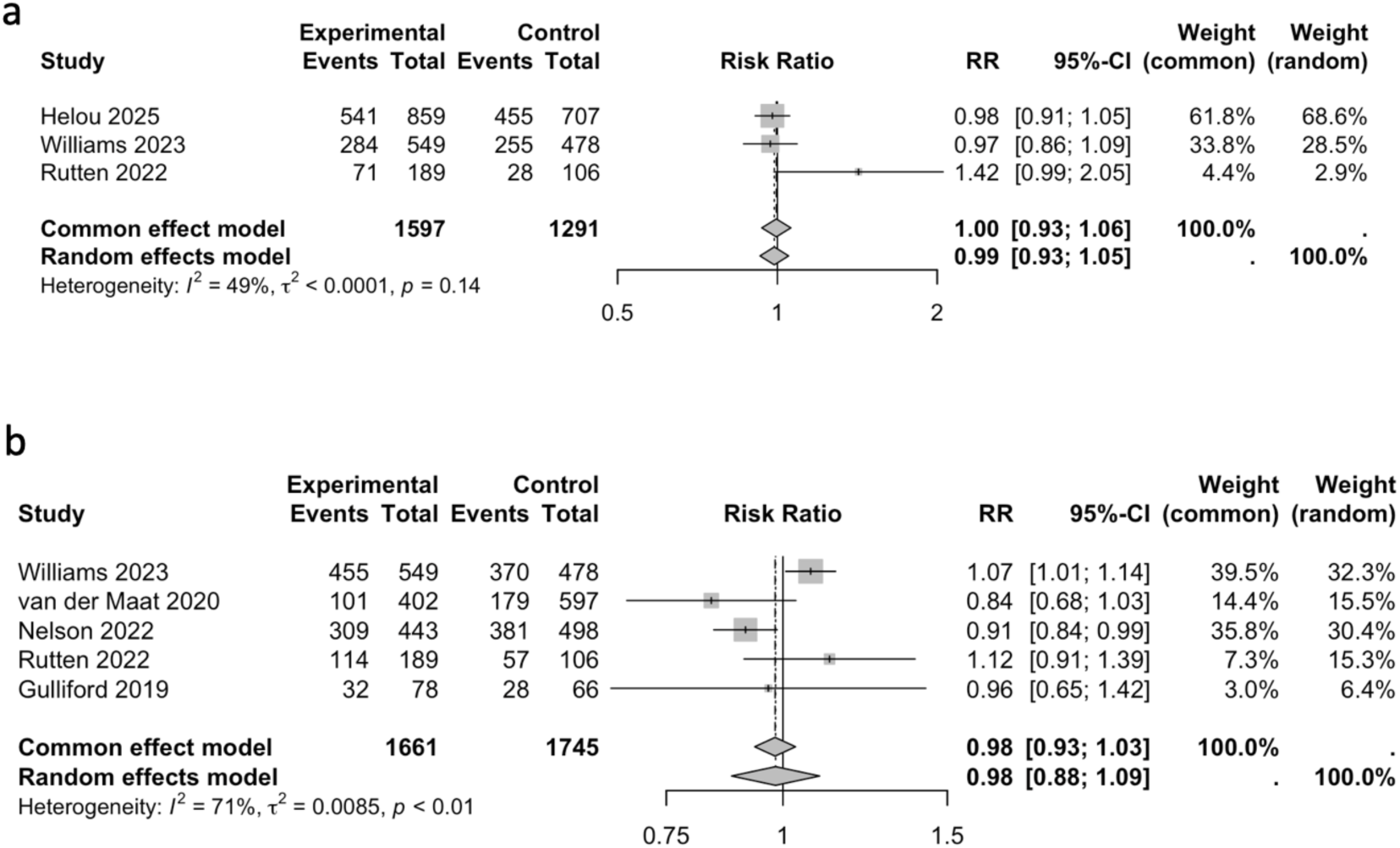
Effect of digital interventions for antimicrobial stewardship on the appropriateness of antibiotic prescription (a) and antibiotic prescription rate (b). RR: Relative Risk; 95% CI, 95% confidence interval.

**Figure 4:**
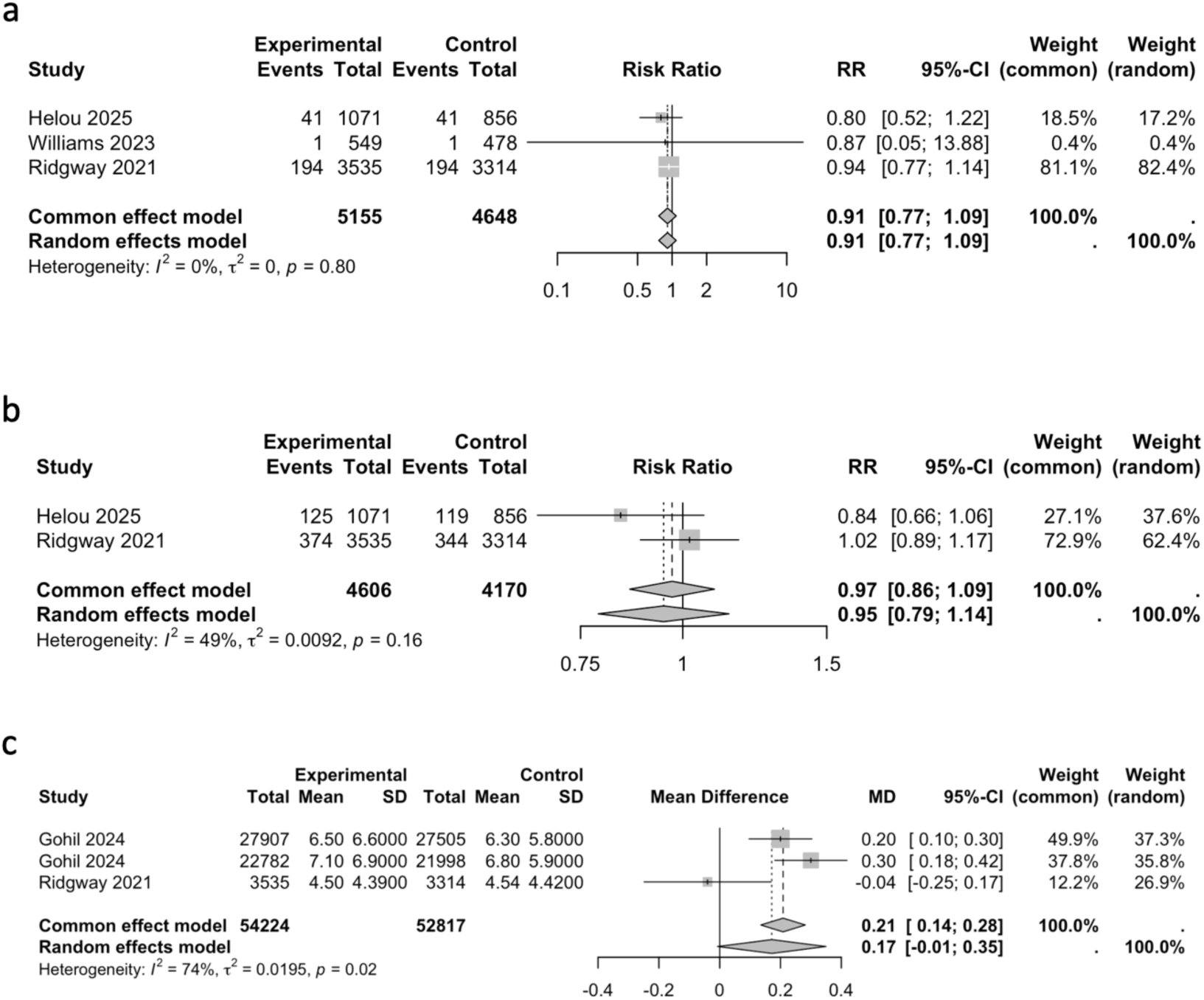
Effect of digital interventions for antimicrobial stewardship on the 30-day mortality (a), 30-day hospital readmission (b), and length of hospital stay (LOS) (c). RR: Relative Risk; MD: Mean Difference; 95% CI, 95% confidence interval.

### Secondary outcomes

The pooled rate did not indicate a difference in mortality within 30 days (RR: 0.91, 95%CI: 0.77 - 1.09; p-value = 0.81; I² = 0%) and rate of hospital readmission (RR: 0.95, 95%CI: 0.79 - 1.14; p-value = 0.16; I² = 49%) between groups (Figure 4A and 4B). Moreover, the pooled data suggested a 17% increase in LOS with the use of digital intervention, but the 95% confidence interval crossed the null (MD: 0.17; 95%CI −0.01 to 0.35). (Figure 4C).

Although heterogeneity was very high, we did not perform subgroup analysis, as stratification would yield too few studies per group, and thus pooled results despite the high heterogeneity.

Finally, we prepared a Summary of Findings from RCTs (Table S2) using the GRADE approach to summarize the magnitude of effects of digital interventions for antimicrobial stewardship, the available body of evidence, and the certainty of the evidence for each outcome.

## Discussion

This systematic review and meta-analysis evaluated the efficacy of digital antimicrobial stewardship interventions in randomized controlled trials. Overall, we found no statistically significant benefits for key outcomes: 30-day mortality, readmissions, antimicrobial prescription rate, and appropriateness. While the point estimate indicated a 17% increase in LOS with the use of digital intervention, the results were imprecise and compatible with no effect. This finding warrants careful interpretation and emphasizes the need to reassess how these tools are designed, implemented, and evaluated. The evidence base was predominantly hospital-based, with only one study conducted in primary care, one in a nursing home facility, and none specifically designed for resource-limited or remote settings lacking on-site infectious diseases expertise. This gap is particularly concerning given that a systematic review published in 2022 highlighted that only 26% of LMIC hospitals have formalized AMS programs, with the scarcity of infectious diseases expertise cited as the foremost barrier, leading to reliance on suboptimal prescribing and heightened AMR risks in resource-constrained environments.^23^

The eleven trials evaluated a diverse range of digital tools, most often clinical decision support systems (CDSS) integrated with electronic health records (EHR) or used as standalone systems, along with order sets, prediction algorithms, and feedback on prescribing. Many of these tools use simple rule-based designs that interrupt workflows and do not fully consider the patient’s specific situation, local infection patterns, or how clinicians work, which can increase mental workload and lead to low use or alert fatigue, a well-documented challenge with rule-based CDSS.^24^ To improve future digital AMS tools, it is imperative to focus on designing them with users in mind, better integration with workflows and EHR systems, and more practical recommendations tailored to the context, such as linking to the reason for treatment, kidney function, allergies, past microbiology results, and previous antibiotic use, while reducing unnecessary alerts.^25^ Importantly, only a single trial assessed long-term effects on antimicrobial resistance, and another one conducted cost analyses, which limits understanding of broader population impacts. Whether these tools can be scaled up affordably is also highlighted in a recent qualitative synthesis of systematic reviews on digital AMS interventions. ^26^

Key limitations of the included RCTs make it harder to draw firm conclusions. The trials differed greatly in their design elements, the intensity of their implementation, ranging from providing simple information to interrupting workflows with prompts, and the degree to which they were carried out, which reduces the comparability of the results and may weaken the overall findings. Most studies were conducted in a single hospital and in well-equipped settings, which limits their relevance to settings with limited technology. Although three trials were conducted in primary care, two were excluded from the meta-analysis for the reasons previously described, thereby limiting the generalizability of the results to this setting. Common methodological problems included risks of bias, incomplete details on other interventions happening at the same time, and varying definitions of what counts as “appropriate” antimicrobial use. Many studies were too small to reliably detect effects on patient outcomes and did not evaluate how well the tools worked in practice, such as ease of use, acceptance by users, alert overload, and how often recommendations were followed, which are crucial for understanding why some results were neutral or showed unintended effects, as well as longer hospital stays. Collectively, these limitations resulted in a very low certainty of evidence across all outcomes according to the GRADE approach. Most included trials had “some concerns” of bias, primarily related to deviations from intended interventions, and the small number of studies per outcome contributed to imprecision bias. Another limitation is that cluster-randomized RCTs lacked adjustment for clustering. This potentially biased results toward significance. No LOS medians were reported, so we used the mean difference (MD) assuming normality. As a result, the true effects of digital antimicrobial stewardship interventions remain uncertain, and the observed estimates should be interpreted with considerable caution.

Our results align with and extend prior syntheses in this field. A qualitative synthesis of systematic reviews of hospital-based digital AMS interventions reported improvements in antimicrobial use and prescribing appropriateness but underscored heterogeneity and limited evidence for patient-centered outcomes.^26^ A systematic review and perspective on telehealth in AMS published in 2021 highlighted the promise of remote stewardship support and audit-and-feedback models, particularly where expertise is limited.^27^ However, although earlier reviews suggested reductions in antimicrobial use, their scope was largely confined to non-randomized studies published up to 2020, limiting causal inference and leaving uncertainty about clinical outcomes. Consequently, the literature has lacked a comprehensive synthesis focused on more recent RCTs assessing the impact of digital interventions on AMS outcomes. Our review addresses this gap; however, the evidence showed no significant impact on downstream outcomes (clinical outcomes, LOS, mortality), suggesting it may not be sufficient to support implementation decisions.

These findings are especially important for remote areas, where effective AMS is hindered by poor access to information and expert teams.^23^ Clinicians often lack timely access to key data needed for good antimicrobial decisions, such as past lab results, local patterns of bacterial resistance, recent antibiotic use, kidney function trends, and allergy records.^28^ Even when digital systems are available, issues such as poor system compatibility, unreliable internet, and incomplete data can make decisions even harder.^29^ At the same time, AMS experts, such as infectious disease doctors, microbiologists, and clinical pharmacists, are usually based in big city hospitals, leaving rural clinics and smaller facilities without ongoing support for reviewing prescriptions, following guidelines, or discussing complex cases. These problems are often worse in LMICs, where limited testing tools and unstable drug supplies lead to overuse of broad-spectrum antibiotics and delays in switching to safer options.^30^ Overall, these barriers highlight the need for digital and telehealth tools that not only provide decision support but also connect remote clinicians with specialists, extending expert AMS functions to places where traditional teams cannot be present.^1,27,31^ Debate persists on the RCT suitability for digital health interventions, which are behavioral by nature, meaning their efficacy hinges on clinician behavior, workflow integration, implementation fidelity, adoption rates, and innovation fatigue. Included studies omitted adoption metrics (e.g., recommendation adherence), risking premature inefficacy conclusions.^32^

Despite limitations of a small, heterogeneous set of RCTs, this review’s key strengths include its focus on randomized evidence, emphasis on critical clinical outcomes (e.g., LOS, mortality), and identification of research gaps in digital AMS evaluation, particularly the need for advanced frameworks such as RE-AIM or MRC guidance and expansion to primary care, resource-limited, and remote settings.^33^

In conclusion, high heterogeneity in interventions and designs yielded inconclusive RCT evidence (high certainty) for digital AMS effects on prescribing and clinical outcomes. RCTs prove suboptimal for behavioral digital health tools, where efficacy depends on adoption and fidelity metrics. Prioritize context-fit AMS platforms for resource-limited/remote settings: user-centered co-design, offline-first/low-bandwidth infrastructure, local-aligned decision support minimizing alert fatigue, plus remote prospective audit via asynchronous review, targeted nudges, and off-site ID escalation for equity. Embed evaluations of processes, adherence, usability, resistance trends, and cost-effectiveness to enable safe scale-up and sustained optimization in underserved contexts.

## Data Availability

All data produced in the present work are contained in the manuscript

## Acknowledgements and funding

This research was funded in part by the Wellcome Trust CAMO-Net programme [grant ref: 226690/Z/22/Z]. For the purpose of open access, the author has applied a CC BY public copyright licence to any Author Accepted Manuscript version arising from this submission. This researcher is affiliated with the Wellcome Trust-funded programme CAMO-Net [grant ref: 226693/Z22/Z].

## Competing interests

All authors declared no competing interests.

## Notes

### Competing Interest Statement

The authors have declared no competing interest.

