## Supplemental material for "Promise vs. Proof in Digital Interventions for Antimicrobial Stewardship: A Systematic Review and Meta-Analysis of Randomized Controlled Trials"


| | |
|---|---|
| Appendix 1 | Pages 2-3 |
| Appendix 2 | Page 4 |
| Table S1 | Pages 5-9 |
| Table S2 | Pages 10-11 |
| Appendix 3 | Page 12 |
| Appendix 4 | Pages 13-15 |

**Appendix 1. Search Strategy**

**Pubmed**

Date: August 22nd, 2025

No date of publication or Language restriction

("digital tool"[Title/Abstract] OR "Computer"[Title/Abstract] OR "Mobile"[Title/Abstract] OR "Electronic"[Title/Abstract] OR "eHealth"[Title/Abstract] OR "Telemedicine"[Title/Abstract]) AND ("Antimicrobial Stewardship"[All Fields] OR "Antibiotic Stewardship"[All Fields] OR "stewardship antibiotic"[All Fields] OR "stewardship antimicrobial"[All Fields])

Records: 1179 results

**BVS / LILACS**

Date: August 22nd, 2025

No date of publication or Language restriction

("digital tool" OR computer OR mobile OR electronic OR ehealth OR telemedicine) AND ("Antimicrobial Stewardship" OR "Antibiotic Stewardship" OR "Stewardship, Antibiotic" OR "Stewardship, Antimicrobial" ) AND instance:"lilacsplus"

Records: 41 results

**Scopus**

Date: August 22nd, 2025

No date of publication or Language restriction

( TITLE-ABS-KEY ( "digital tool" OR Computer OR Mobile OR Electronic OR eHealth OR Telemedicine ) AND TITLE-ABS-KEY ( "Antimicrobial Stewardship" OR "Antibiotic Stewardship" OR "Stewardship, Antibiotic" OR "Stewardship, Antimicrobial" ) )

Records: 2024 results

**Embase**

Date: August 22nd, 2025

No date of publication or Language restriction

('digital tool':ti,ab,kw OR computer:ti,ab,kw OR mobile:ti,ab,kw OR electronic:ti,ab,kw OR ehealth:ti,ab,kw OR telemedicine:ti,ab,kw) AND ('antimicrobial stewardship':ti,ab,kw OR 'antibiotic stewardship':ti,ab,kw OR 'stewardship, antibiotic':ti,ab,kw OR 'stewardship, antimicrobial':ti,ab,kw)

Records: 2042 results

**Appendix 2. Primary outcomes definition according to the included studies for the meta-analysis**

***Appropriateness of antibiotic prescribing***

**Helou (2025):** appropriate antibiotic therapy (AAT) described as adequate selection of drug, route and dose of antibiotic treatments in concordance with local guidelines at the patient level as described before according to a previous study (Schuts EC, et al. Current evidence on hospital antimicrobial stewardship objectives: a systematic review and meta-analysis. Lancet Infect Dis 2016;16:847e56. https://doi.org/10.1016/s1473-3099(16)00065-7).

**Williams (2023):** the primary outcome was exclusive guideline-concordant antibiotic prescribing during the first 24 hours of care. Concordance was pre-specified and emphasized use of narrow-spectrum antibiotics. Withholding antibiotics was presumed to be concordant because childhood pneumonia is frequently viral,4,27 and clinicians were considered highly unlikely to withhold antibiotics in those with suspected bacterial pneumonia.

**Rutten (2022**): the primary study outcome was the percentage of antibiotic prescriptions for suspected UTI that were appropriate (yes/no), that is, prescribed in compliance with the treatment advice generated by the decision tool.

***Antibiotic prescription***

**Williams (2023):** antibiotic prescription per ED encounter during the first 24 hours of care.

**Van der Maat (2020):** antibiotic prescription at ED discharge (yes/no)

**Nelson (2022):** the proportion of children prescribed an antibiotic by the treating physician.

**Gulliford (2019):** antibiotic prescription rate per respiratory tract infection consultation

**Rutten (2022):** total antibiotic prescribing

**Table S1.** Summary of all randomized clinical trials evaluating digital interventions for antimicrobial stewardship

| Author, year | Country | Setting | Patient population | Digital tool studied | Infection site | Control group | Multimodal strategy | Primary Outcomes | Secondary Outcomes | Follow-up assessment | Sample size |
|---|---|---|---|---|---|---|---|---|---|---|---|
| Helou, 2025 | Netherlands, Sweden, Switzerland | Tertiary hospitals | Adult (≥18 years) | Mobile smartphone application / Clinical Decision Support System (CDSS) | Unspecified | Standard care with conventional access to antibiotic guidelines via: website, PDF files, printed booklets | Alone | **Appropriate antibiotic therapy (AAT) rate** | **Length of stay (LOS); mortality 30 days; ICU admission; readmission within 30 days;** Medication-specific usage patterns; user analytics of the app; AAT assessed by user analytics | 16 months | 1,566 |
| Gohil, 2024 | USA | Community hospitals | Adult (≥18 years) | Computerized Provider Order Entry (CPOE) bundle with real-time prompts recommending empiric standard-spectrum antibiotics | Urinary tract infection | Routine stewardship (education + coaching calls) without CPOE prompts; patient-specific risk estimates calculated but not displayed | Multimodal (Real-time CPOE prompts for MDRO risk <10%; patient-specific risk estimates; educational materials; feedback reports; coaching calls; site visits & webinars | Empiric extended-spectrum antibiotic days of therapy (first 3 hospitalization days) | Vancomycin days of therapy; antipseudomonal days of therapy; days to antibiotic escalation; days to ICU transfer; length of stay in days | 15 months | 55,412 |
| Gohil, 2024 | USA | Community hospitals | Adult (≥18 years) | Computerized Provider Order Entry (CPOE) bundle with real-time prompts recommending empiric standard-spectrum antibiotics | Respiratory tract infections | Routine stewardship (education + coaching calls) without CPOE prompts; patient-specific risk estimates calculated but not displayed | Multimodal (Real-time CPOE prompts for MDRO risk <10%; patient-specific risk estimates; educational materials; feedback reports; coaching calls; site visits & webinars | Empiric extended-spectrum antibiotic days of therapy (first 3 hospitalization days) | Vancomycin days of therapy; antipseudomonal days of therapy; days to antibiotic escalation; days to ICU transfer; length of stay in days | 15 months | 44,780 |

| | | | | | | | | | | | |
|---|---|---|---|---|---|---|---|---|---|---|---|
| Williams, 2023 | USA | Pediatric emergency departmen ts | Pediatric (6 months to < 18 years) | EHR-based antibiotic CDS in the ED | Respiratory tract infections | Standard care with access to local guidelines, but no CDS tool | Multimodal (EHR screening for pneumonia, CDS alerts with tailored regimens, education sessions, emails, and ED clinician champions) | Exclusive guideline-concordant antibiotic prescribing within the first 24 hours of care | Any guideline-concordant prescribing (24 h and entire encounter); safety (time to first antibiotic, LOS, delayed ICU transfer, 3- and 7-day revisits, 30-day mortality) | First 24 h of care, entire encounter, 3- and 7-day post-discharge revisits, and 30-day mortality | 1,027 |
| Van der Maat, 2020 | Netherland s | Emergency departmen ts | Pediatric (1 to 60 months) | Feverkidstool: Online digital calculator that stratifies fever-related infection risk using temperature, CRP, and clinical appearance, requiring manual access by clinicians before treatment decisions | Respiratory tract infections | Standard care (without CDS tool) | Bimodal (Fevekidstool + education) | Overall antibiotic prescription (binary); Strategy failure: antibiotic use + hospitalization + oxygen dependency or fever up to day 7 + complications | Compliance (intention-to-treat + per-protocol analyses); number of complications | Primary outcome at ED discharge; Strategy failure assessed at day 7 follow-up | 999 |
| Nelson, 2022 | Bangladesh and Mali | Governmen t hospitals and healthcare facilities | Pediatric (2 to 59 months) | Smartphone-based electronic Clinical Decision Support (eCDS) with Diarrheal Etiology Prediction (DEP) algorithm | Acute diarrhea | eCDS without DEP algorithm (standard smartphone eCDS); no algorithm-based prediction support | Alone | Proportion of children prescribed antibiotics | Diarrheal symptom resolution at 10 days post-discharge | Baseline (enrollment); 4-week intervention period; 10 days post-discharge follow-up | 941 |
| Rutten, 2022 | Netherland s | Nursing Homes | Older adults NH residents (mean age 86 years) | An electronic health record (EHR)-integrated decision tool, based on a previously developed decision tool for the | Urinary tract infection | Standard care | Multimodal (CDS tool + supportive interventions: interactive training session proprovided by | Percentage of antibiotic prescriptions for suspected UTI that were appropriate (in compliance with the | Changes in treatment decision (ie, antibiotic start after initial withholding antibiotics); complications (side effects, renal | 3, 7, and 21 days later in the intervention group; 7 and 21 | 212 |

| | | | | | | | | | | | |
|---|---|---|---|---|---|---|---|---|---|---|---|
| | | | | treatment of suspected UTI in frail older adults | | | the research team, pocket-cards, and an information leaflet to hand out to residents) | treatment advice generated by the decision tool) | impairment, and pyelonephritis/urosepsis); UTI-related hospitalization; mortality during follow-up, and pre-post study changes in total antibiotic prescribing | days later in the control group | |
| Ridgway, 2021 | USA | Univeristy hospitals (Healthsystem) | Adult (≥18 years) | Electronic Clinical Decision Support system (CDSS) (Weighted Incidence Syndromic Combination Antibiogram (WISCA)) | Urinary tract infection; abdominal biliary infection; nonpurulent cellulitis; community acquired pneumonia; aspiration pneumonia, and nursing home-associated pneumonia. | Standard care with audit and feedback recorded in EHR but not communicated to providers unless the antibiotic regimen posed a clinical threat; no active prospective feedback intervention | Multimodal (EHR-based CDS tool + guideline-based empiric regimens + prospective audit and feedback by ASP physicians | Hospital length of stay (LOS) | 30-day readmission; 30-day mortality; Antibiotic charges; Clostridioides difficile infection acquisition within 180 days; Multidrug-resistant gram-negative organism (MDRO) acquisition within 180 days | In-hospital, 30-day, and 180-day follow-up | 6,849 |
| Gulliford, 2019 | United Kingdom | Primary care | General population | Clinical Practice Research Datalink (CPRD) integrated decision support | Respiratory tract infections | Standard care (usual prescribing practices; access to routine resources) | Multimodal (Webinar + Monthly data reports + Decision support tools (not specified if mobile/web) | Antibiotic prescription rate for RTI per 1,000 patient-years (12 months) | RTI consultation rate; Proportion of RTI consultations with antibiotics prescribed; Total antibiotic-prescribing rate; Subgroups of RTI; Health-care costs | 12 months | 626,625 |

| | | | | | | | | | | | |
|---|---|---|---|---|---|---|---|---|---|---|---|
| Broussard, 2025[1] | USA | Outpatient pediatric primary care clinics | Pediatric (3 months to <21 years) | An HER SSTI order panel with default, pre-populated guideline-concordant antibiotic durations and weight-based dosing | Skin and soft tissue infections | Control EHR SSTI order panel listing the same antibiotics but without any suggested durations or pre-populated dosing | Educational sessions on SSTI treatment guidelines + SSTI EHR order panel with default durations and weight-based dosing + visual reminders near workstations + reminder email about panel use | Inappropriately long antibiotic prescriptions rate for SSTIs (guidelines-based cutoffs) | Change in trajectories (pre- vs post-panel) of long prescription; rates over time; odds of long prescriptions; frequency of order panel use | 12 months | 1,123 |
| Yan, 2020[2] | USA | Direct-to-Patient Telemedicine | General population | HER-integrated Audit & Feedback via Online Personalized Dashboard (web-based) | Respiratory tract infections | Standard Care (Education Only) | Education + Online feedback dashboard with: Practice Summary (aggregate rates), Individual clinician rates + Digital Monthly personalized dashboard | Antibiotic prescription rates for URI, bronchitis, sinusitis, pharyngitis; | Diagnostic shifting: proportion of visits coded as sinusitis/pharyngitis (potentially antibiotic-appropriate vs inappropriate conditions) | 6 months | 24,843 |

[1] Not included for quantitative synthesis (reason for exclusion: assessed different outcomes from those described in the protocol)
[2] Not included for quantitative synthesis (reason for exclusion: missing critical data)

**Table S2.** GRADE Summary of Findings from Randomized Clinical Trials on Digital Interventions for Antimicrobial Stewardship

| Outcome | Relative effect (95% CI)[a] | No. of studies | Certainty of evidence[b] | Comment |
|---|---|---|---|---|
| Appropriateness of antibiotic prescription | RR 0.99 (0.93 – 1.05) | 3 RCTs | ⊕○○○ Very low | Certainty reduced due to serious risks of bias in study design, inconsistency in the direction of effects across the three trials, and significant imprecision due to the small cumulative sample size. |
| Antibiotic prescription | RR 0.98 (0.88 – 1.09) | 5 RCTs | ⊕○○○ Very low | Certainty downgraded for risk of bias (predominantly deviations from intended interventions), serious inconsistency (substantial heterogeneity, $I^2$=71%, with effects in opposite directions), imprecision (pooled CI crossing the null), and suspected publication bias (only five studies). |
| 30-day mortality | RR 0.91 (0.77 – 1.09) | 3 RCTs | ⊕○○○ Very low | Certainty downgraded for risk of bias (concerns in deviations from intended interventions, including one high-risk trial), imprecision (pooled CI crossing the null), and suspected publication bias (only three studies). |
| 30-day hospital readmission | RR 0.95 (0.79 – 1.14) | 3 RCTs | ⊕○○○ Very low | Certainty downgraded for risk of bias (including one high-risk trial, primarily due to deviations from intended interventions), inconsistency (heterogeneity and opposing point estimates across two studies), imprecision (pooled CI crossing the null), and suspected publication bias (only two studies). |
| Length of hospital stay (LOS) | MD 0.17 (-0.01 – 0.35) | 3 RCTs | ⊕○○○ Very low | Certainty downgraded for risk of bias (including one high-risk trial), serious inconsistency (substantial heterogeneity, $I^2$=74%, with effects in opposite directions), imprecision (pooled CI crossing the null), and suspected publication bias (only three studies). |

Abbreviations: RCTs, randomized controlled trials.

[a] RR (Relative Risk) or MD (Mean Difference) with 95% confidence interval obtained in the meta-analyses.

[b] GRADE Working Group grades of evidence: High = This research provides a very good indication of the likely effect. The likelihood that the effect will be substantially different* is low. Moderate = This research provides a good indication of the likely effect. The likelihood that the effect will be substantially different* is moderate. Low = This research provides some indication of the likely effect. However, the likelihood that it will be substantially different* is high. Very low = This research does not provide a reliable indication of the likely effect. The likelihood that the effect will be substantially different* is very high. *Substantially different = a large enough difference that it might affect a decision.

**Appendix 3.** PRISMA 2020 for Abstracts Checklist

| Section and Topic | Item # | Checklist item | Reported (Yes/No) |
|---|---|---|---|
| **TITLE** | | | |
| Title | 1 | Identify the report as a systematic review. | Yes |
| **BACKGROUND** | | | |
| Objectives | 2 | Provide an explicit statement of the main objective(s) or question(s) the review addresses. | Yes |
| **METHODS** | | | |
| Eligibility criteria | 3 | Specify the inclusion and exclusion criteria for the review. | Yes |
| Information sources | 4 | Specify the information sources (e.g. databases, registers) used to identify studies and the date when each was last searched. | Yes |
| Risk of bias | 5 | Specify the methods used to assess risk of bias in the included studies. | Yes |
| Synthesis of results | 6 | Specify the methods used to present and synthesise results. | Yes |
| **RESULTS** | | | |
| Included studies | 7 | Give the total number of included studies and participants and summarise relevant characteristics of studies. | Yes |
| Synthesis of results | 8 | Present results for main outcomes, preferably indicating the number of included studies and participants for each. If meta-analysis was done, report the summary estimate and confidence/credible interval. If comparing groups, indicate the direction of the effect (i.e. which group is favoured). | Yes |
| **DISCUSSION** | | | |
| Limitations of evidence | 9 | Provide a brief summary of the limitations of the evidence included in the review (e.g. study risk of bias, inconsistency and imprecision). | Yes |
| Interpretation | 10 | Provide a general interpretation of the results and important implications. | Yes |
| **OTHER** | | | |
| Funding | 11 | Specify the primary source of funding for the review. | Yes |
| Registration | 12 | Provide the register name and registration number. | Yes |

*From:* Page MJ, McKenzie JE, Bossuyt PM, Boutron I, Hoffmann TC, Mulrow CD, et al. The PRISMA 2020 statement: an updated guideline for reporting systematic reviews. BMJ 2021;372:n71. doi: 10.1136/bmj.n71. This work is licensed under CC BY 4.0. To view a copy of this license, visit https://creativecommons.org/licenses/by/4.0/

**Appendix 4.** PRISMA 2020 Checklist

| Section and Topic | Item # | Checklist item | Location where item is reported |
|---|---|---|---|
| **TITLE** | | | |
| Title | 1 | Identify the report as a systematic review. | 1-2 |
| **ABSTRACT** | | | |
| Abstract | 2 | See the PRISMA 2020 for Abstracts checklist. | 28-61 |
| **INTRODUCTION** | | | |
| Rationale | 3 | Describe the rationale for the review in the context of existing knowledge. | 77-96 |
| Objectives | 4 | Provide an explicit statement of the objective(s) or question(s) the review addresses. | 97-105 |
| **METHODS** | | | |
| Eligibility criteria | 5 | Specify the inclusion and exclusion criteria for the review and how studies were grouped for the syntheses. | 130-142 |
| Information sources | 6 | Specify all databases, registers, websites, organisations, reference lists and other sources searched or consulted to identify studies. Specify the date when each source was last searched or consulted. | 115-118 |
| Search strategy | 7 | Present the full search strategies for all databases, registers and websites, including any filters and limits used. | 119-128; Supplementary material (Appendix 1) |
| Selection process | 8 | Specify the methods used to decide whether a study met the inclusion criteria of the review, including how many reviewers screened each record and each report retrieved, whether they worked independently, and if applicable, details of automation tools used in the process. | 142-145 |
| Data collection process | 9 | Specify the methods used to collect data from reports, including how many reviewers collected data from each report, whether they worked independently, any processes for obtaining or confirming data from study investigators, and if applicable, details of automation tools used in the process. | 147-154 |
| Data items | 10a | List and define all outcomes for which data were sought. Specify whether all results that were compatible with each outcome domain in each study were sought (e.g. for all measures, time points, analyses), and if not, the methods used to decide which results to collect. | 155-161; Supplementary material (Appendix 2) |
| | 10b | List and define all other variables for which data were sought (e.g. participant and intervention characteristics, funding sources). Describe any assumptions made about any missing or unclear information. | Supplementary material (Table S1) |
| Study risk of bias assessment | 11 | Specify the methods used to assess risk of bias in the included studies, including details of the tool(s) used, how many reviewers assessed each study and whether they worked independently, and if applicable, details of automation tools used in the process. | 163-165 |

| Section and Topic | Item # | Checklist item | Location where item is reported |
|---|---|---|---|
| Effect measures | 12 | Specify for each outcome the effect measure(s) (e.g. risk ratio, mean difference) used in the synthesis or presentation of results. | 171-177 |
| Synthesis methods | 13a | Describe the processes used to decide which studies were eligible for each synthesis (e.g. tabulating the study intervention characteristics and comparing against the planned groups for each synthesis (item #5)). | 130-142; Supplementary material (Table S1) |
| | 13b | Describe any methods required to prepare the data for presentation or synthesis, such as handling of missing summary statistics, or data conversions. | 159-161; 201-205 |
| | 13c | Describe any methods used to tabulate or visually display results of individual studies and syntheses. | Supplementary material (Table S1); 247-287 |
| | 13d | Describe any methods used to synthesize results and provide a rationale for the choice(s). If meta-analysis was performed, describe the model(s), method(s) to identify the presence and extent of statistical heterogeneity, and software package(s) used. | 171-177; 281-283 |
| | 13e | Describe any methods used to explore possible causes of heterogeneity among study results (e.g. subgroup analysis, meta-regression). | NA |
| | 13f | Describe any sensitivity analyses conducted to assess robustness of the synthesized results. | NA |
| Reporting bias assessment | 14 | Describe any methods used to assess risk of bias due to missing results in a synthesis (arising from reporting biases). | 247-254 |
| Certainty assessment | 15 | Describe any methods used to assess certainty (or confidence) in the body of evidence for an outcome. | 284-287 Supplementary material (Table S2); |
| **RESULTS** | | | |
| Study selection | 16a | Describe the results of the search and selection process, from the number of records identified in the search to the number of studies included in the review, ideally using a flow diagram. | 208-245; |
| | 16b | Cite studies that might appear to meet the inclusion criteria, but which were excluded, and explain why they were excluded. | 182-187; 208-245; 201-202 |
| Study characteristics | 17 | Cite each included study and present its characteristics. | 186-187; Supplementary material (Table S1); |
| Risk of bias in studies | 18 | Present assessments of risk of bias for each included study. | 247-251 |

| Section and Topic | Item # | Checklist item | Location where item is reported |
|---|---|---|---|
| Results of individual studies | 19 | For all outcomes, present, for each study: (a) summary statistics for each group (where appropriate) and (b) an effect estimate and its precision (e.g. confidence/credible interval), ideally using structured tables or plots. | 256-281 |
| Results of syntheses | 20a | For each synthesis, briefly summarise the characteristics and risk of bias among contributing studies. | 247-251; 256-281 |
| | 20b | Present results of all statistical syntheses conducted. If meta-analysis was done, present for each the summary estimate and its precision (e.g. confidence/credible interval) and measures of statistical heterogeneity. If comparing groups, describe the direction of the effect. | 256-281 |
| | 20c | Present results of all investigations of possible causes of heterogeneity among study results. | NA |
| | 20d | Present results of all sensitivity analyses conducted to assess the robustness of the synthesized results. | NA |
| Reporting biases | 21 | Present assessments of risk of bias due to missing results (arising from reporting biases) for each synthesis assessed. | NA |
| Certainty of evidence | 22 | Present assessments of certainty (or confidence) in the body of evidence for each outcome assessed. | 284-287; Supplementary material (Table S2); |
| **DISCUSSION** | | | |
| Discussion | 23a | Provide a general interpretation of the results in the context of other evidence. | 292-321 |
| | 23b | Discuss any limitations of the evidence included in the review. | 322-340 |
| | 23c | Discuss any limitations of the review processes used. | 340-345 |
| | 23d | Discuss implications of the results for practice, policy, and future research. | 346-393 |
| **OTHER INFORMATION** | | | |
| Registration and protocol | 24a | Provide registration information for the review, including register name and registration number, or state that the review was not registered. | 112-113 |
| | 24b | Indicate where the review protocol can be accessed, or state that a protocol was not prepared. | 112-113 |
| | 24c | Describe and explain any amendments to information provided at registration or in the protocol. | NA |
| Support | 25 | Describe sources of financial or non-financial support for the review, and the role of the funders or sponsors in the review. | 396-403 |
| Competing interests | 26 | Declare any competing interests of review authors. | 406-408; ICMJE form |
| Availability of data, code and other materials | 27 | Report which of the following are publicly available and where they can be found: template data collection forms; data extracted from included studies; data used for all analyses; analytic code; any other materials used in the review. | NA |

*From:* Page MJ, McKenzie JE, Bossuyt PM, Boutron I, Hoffmann TC, Mulrow CD, et al. The PRISMA 2020 statement: an updated guideline for reporting systematic reviews. BMJ 2021;372:n71. doi: 10.1136/bmj.n71. This work is licensed under CC BY 4.0. To view a copy of this license, visit https://creativecommons.org/licenses/by/4.0/
